# Comprehension in chronic aphasia relies on the language network, not the Multiple Demand network

**DOI:** 10.64898/2026.03.16.26348460

**Authors:** Anne Billot, Maria Varkanitsa, Niharika Jhingan, Nicole Carvalho, Isaac Falconer, Hannah Small, Rachel Ryskin, Idan Blank, Evelina Fedorenko, Swathi Kiran

## Abstract

Post-stroke aphasia recovery has been proposed to be supported by two broad mechanisms: i) reorganization within the remaining components of the language network; and ii) recruitment of brain areas outside of the premorbid language network to assume language function post stroke. A key candidate for such network reorganization is the multiple demand (MD) network, which supports executive functions and is recruited for a wide range of cognitive tasks. A prominent hypothesis suggests that, when specialized language resources are no longer sufficient, this network supports linguistic processing while retaining its domain-general role.

In this cross-sectional study, we used precision functional MRI to evaluate these two hypotheses in 37 individuals with chronic post-stroke aphasia, along with 38 age-matched controls. Language and MD networks were identified within individuals using functional localizer tasks. Participants with aphasia completed extensive behavioral assessments that evaluated linguistic and general cognitive skills. We examined responses during language processing in each of the two networks and related activity and functional connectivity measures from the two networks to linguistic ability.

Our results do not support the hypothesis that language comprehension in chronic aphasia recruits parts of the MD network. First, the language and the MD networks remain robustly dissociated: the language network responds selectively to language across modalities, and no MD region shows increased responses to language comprehension in aphasia relative to controls. Second, functional connectivity analyses reveal no increased integration between the two networks during language comprehension. Third, linguistic ability is associated with the strength of activity within the language network, but not the MD network.

Although we cannot rule out a role for the MD network in aphasia recovery during the acute or subacute phases, or in more severely impaired patients, it appears that during the chronic phase, language comprehension relies on the same specialized network as in healthy controls.

## Introduction

Aphasia affects over a third of stroke survivors^1^, and the incidence of stroke continues to rise as the aging population increases^2^. Aphasia severely affects patients’ quality of life^3–5^ and up to 90% of persons with aphasia (PWA) do not fully recover their linguistic abilities one year after stroke^6^. As a result, developing effective treatments remains a core goal in aphasia research^7,8^. The degree of language recovery varies substantially across PWA^9–13^ and depends on a multitude of factors. In particular, the neuroplasticity mechanisms that underlie this variability remain poorly understood (for reviews, see ^14–19)^. The nature of the neural mechanisms supporting language in the chronic stage of recovery remains debated, yet it is critical both for understanding the fundamental principles of neuroplasticity^20–23^ and for informing targeted rehabilitation strategies.

One central unresolved question concerns the degree to which language function following stroke relies on the residual specialized language system or requires broader cortical reorganization of function, including recruitment of other brain networks. The most common approach to probing the neural infrastructure of language processing in aphasia is to examine brain responses (e.g., with functional MRI, fMRI) while PWA perform language tasks. In most neurologically healthy individuals, linguistic processing relies on a left-lateralized fronto-temporal network^24–29^. This network is strongly selective for linguistic inputs (for a review, see ^30^) and supports computations related to word recognition and combinatorial syntactic and semantic processing during both comprehension and production (e.g., ^31–33)^. These language regions emerge in early childhood ^34–40^ and continue to support language processing across the adult lifespan (e.g., ^41–44;^ for reviews, see ^45,46^).

The prevailing hypothesis about aphasia recovery has been that the spared regions of this specialized language network continue to support language processing after stroke (e.g., ^15,17,47–50)^, and that recovery occurs either through homeostatic plasticity or within-network reconfiguration (see e.g.,^14^ for discussion). Indeed, many studies have observed neural responses within this canonical language network in PWA performing language tasks ^19,48,50–57;^ see review by ^15^. Whether, and to what extent, stroke-induced damage to this network also triggers functional reorganization that involves other brain networks—such as homotopic contralesional language regions^15,18,50,53,58–60^ or the domain-general multiple demand (MD) network^15,18,54,55,61–69^—remains debated.

Activation in the right-hemisphere homotopic language regions is perhaps expected given that a) in neurotypical adults, these areas show some engagement during language processing, although the responses are weaker and less selective than in the core left-hemisphere language regions (e.g., ^28,70–75)^, and b) in cases of early left hemisphere damage, the right hemisphere language network can successfully support language function^76–78^. Further, in neurologically healthy individuals, perturbation of activity in the left-hemisphere language regions using brain stimulation leads to increased activity in the right-hemisphere homotopic regions^79,80^. Evidence from aphasia suggests that right hemisphere recruitment is prominent in the acute and subacute phases, but in PWA who recover linguistic abilities well, activity shifts back to the spared left-hemisphere regions over time, with persistent right hemisphere reliance associated with poorer long-term outcomes^50,51,55,81,82^.

The role of the domain-general MD regions in aphasia recovery is more controversial. The MD network—a bilateral network with prominent components in the frontal and parietal cortex—is engaged during diverse demanding cognitive tasks and has been associated with psychological constructs such as working memory, attention, and cognitive control^63,64,83–86^. Individual differences in activation within this network in neurotypical adults have been linked with differences in general fluid intelligence^87,88^, and damage to the MD brain regions is associated with loss of fluid reasoning and executive control abilities^89–94^. Importantly, in healthy brains, this network is distinct from the language network with no response during language processing and strong responses during diverse executive function tasks, and near-zero functional connectivity with the language regions (e.g., ^95–100;^ see ^101,102^ for reviews). Even when language processing gets complex, in the presence of rare words or constructions, or when non-local syntactic dependencies need to be formed, the left-hemisphere language regions, but not the MD network, show a strong response and modulation by linguistic complexity^103–105^ (see ^106^ for review). The MD regions only show engagement—in addition to the language areas—during language tasks where language processing is accompanied by extraneous task demands, such as semantic judgments or comprehension questions about the sentences^99,107,108^. Thus, this body of evidence from neurotypical adults suggests that the MD network does not contribute to linguistic processing beyond being engaged by general task demands in paradigms that include tasks.

However, another important property of the MD network is its functional flexibility; these brain regions have long been implicated in learning new skills and solving new problems ^64,109,110^. This flexibility gave rise to the hypothesis that the MD network can take on new functions—including aspects of language—when specialized processing mechanisms get damaged^17,111,112^. This hypothesis has received support in the form of activation in the PWA’s putative MD areas during language tasks, including in the dorsal anterior cingulate cortex or the inferior and middle frontal gyri^18,54,55,61,65–67,69^. Additional support has come from brain-behavior association studies, including those that rely on lesion-symptom mapping^113^, or static and dynamic functional connectivity^61,65,68,114,115^.

However, the evidence for the role of the MD network in aphasia recovery has been challenged. For example, based on a meta-analysis of past studies, Wilson & Schneck^19^ have argued that the reported upregulation of frontal and parietal regions can be explained in terms of the task demands that accompany most language paradigms used in studies of PWA, in line with fMRI evidence from healthy participants discussed above. Another limitation of prior fMRI studies of PWA is that most have relied on group-level analyses where individual brains are averaged in the common space. Because of the well-established inter-individual differences in functional topography^116–121^, this averaging leads to the blurring of functional network boundaries, which is especially problematic when trying to differentiate between adjacent brain networks, such as the language and the MD networks that lie side by side within the frontal lobe^96,97,102,122^. Indeed, one prior fMRI study that has relied on individual-subject analyses^123^ did not find evidence of the MD network’s engagement during language processing in chronic aphasia.

Here, we adopt the individual-subject precision fMRI approach, relying on previously validated ‘localizers’ for the language and MD networks^85,116,124,125^, to test whether the MD regions, defined functionally within individuals, show greater engagement during language comprehension in individuals with chronic aphasia, relative to age-matched controls. We complement these analyses of the two networks’ activation profiles with a detailed cognitive-behavioral assessment of linguistic and cognitive abilities in PWA to ask which aspects of neural activity best predict linguistic competence (preregistration for this component of the study: https://osf.io/xcfpz/). If the MD network supports linguistic processing in chronic aphasia, we should observe i) MD network engagement during language comprehension tasks, ii) increased integration between the language and the MD networks, as reflected in functional connectivity, and iii) associations between MD network activity or connectivity and linguistic competence. If, instead, the residual language network remains the neural substrate for linguistic processing in PWA, we should observe selective engagement of the language regions during language comprehension tasks, persistent segregation between the two networks in their functional connectivity patterns, and associations between the language network activity/connectivity and linguistic ability. In exploratory analyses, we evaluated whether aphasia severity and lesion location affect the engagement of the two networks during language processing, to test whether greater aphasia severity may be associated with reduced language-network engagement and increased recruitment of the MD network.

## Materials and methods

Additional methods details are provided in the **Supplementary Materials**.

### Participants

37 individuals (seven female; mean age = 58.3 years, SD = 8.4; mean education = 15.6 years, SD = 2.9) who received a diagnosis of aphasia after a single left-hemisphere stroke (mean time post-stroke onset = 84.5 months, SD = 94.1; see **Fig. 1A** for demographics summary and lesion overlap) and 38 healthy controls (21 female; mean age = 61.6 years, SD = 9.2; mean education = 18.5 years, SD = 4.1) were recruited between October 2020 and August 2024. PWA and controls did not differ significantly in age (p = 0.102) but controls had significantly more years of education (p <0.001). Participants with aphasia (PWA) were all in the chronic phase of stroke recovery (> six months post-stroke). Participants were native English speakers (including three bilingual individuals with aphasia; for all three, all the testing was done in English), had normal or corrected-to-normal hearing and vision, and were medically and neurologically stable. Four additional PWA and four controls were excluded from the fMRI analyses for inability to complete tasks in the MRI or because their age fell substantially outside the range of the rest of the cohort. Among the 37 PWA, four completed the behavioral battery more than 9 months from their MRI session because of COVID-19 restrictions and were therefore excluded from the brain-behavior analyses. For the remaining 33 PWA, the mean interval between scanning and behavioral testing was 59 days (SD = 70 days). Three of the four excluded PWA were retained in the analyses of aphasia severity (n = 36), as their WAB-AQ was collected within two months of scanning. Detailed demographic information is available in **Supplementary Table 1**. The mean Western Aphasia Battery-Revised (WAB-R)^126^ aphasia quotient (AQ) across all 37 PWA was 80.6 (SD = 22.1, min = 12.4, max=100). All PWA reported persistent language difficulties in their daily life at the time of the study, although fifteen PWA scored at or above the WAB-R cut-off for recovery (AQ ≥ 93.8), indicating they no longer met diagnostic criteria for aphasia at the time of testing (see **Supplementary Figs. 1** and 2 for evidence that the data are similar when this subset is excluded). All participants gave written informed consent in accordance with the requirements of MIT’s Committee on the Use of Humans as Experimental Subjects (COUHES), the Declaration of Helsinki, and the Institutional Review Board of Boston University. Participants were paid for their participation.

**Figure 1.**
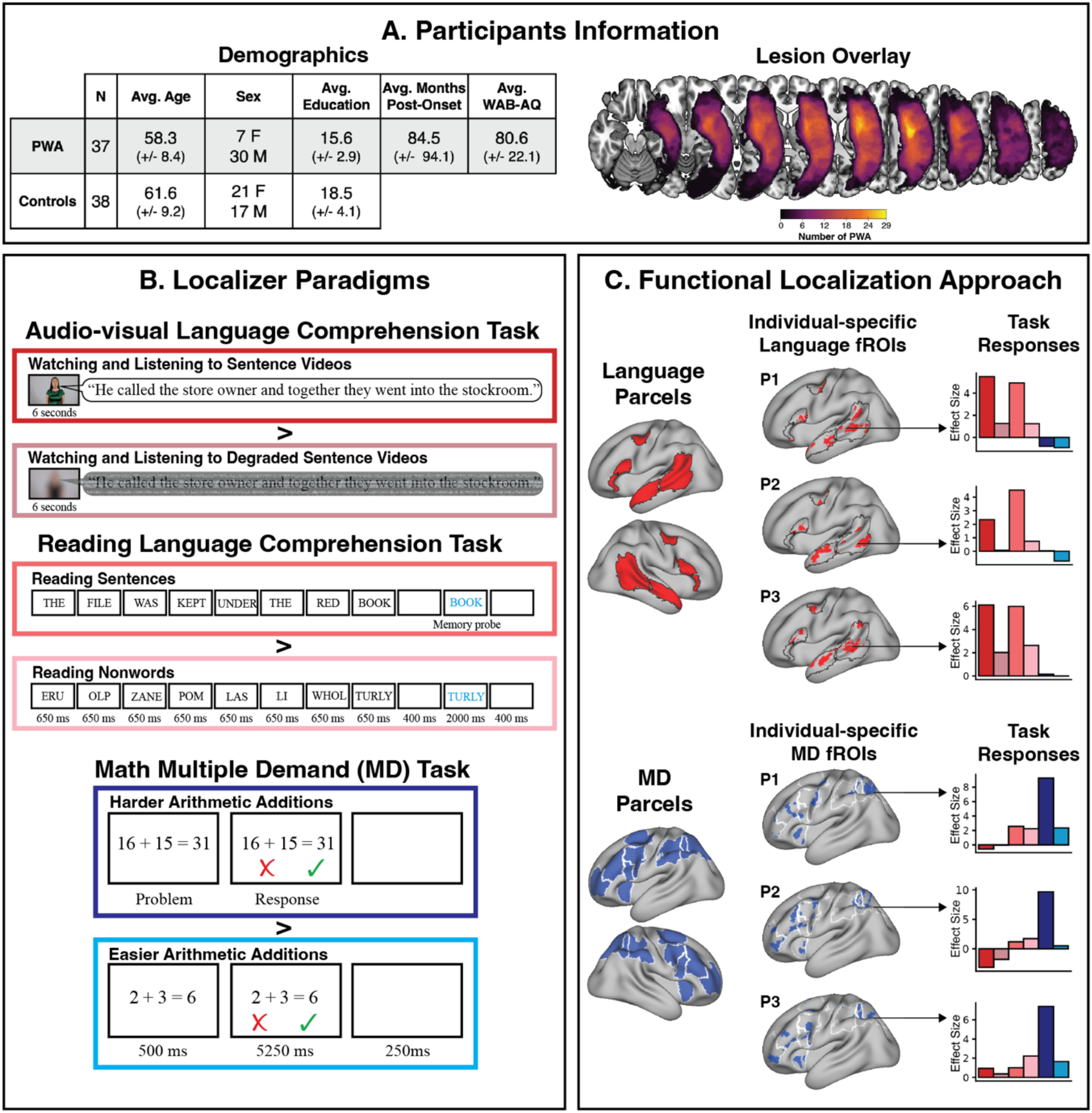
Participants, paradigms, and analytic approach. A/ Lesion overlay and participants’ demographics. B/ Language and MD network localizers used to identify individual-specific language and MD functional regions of interest (fROIs). Two localizers were used for the language network: the audio-visual comprehension task includes blocks where participants watch and listen to sentence videos and control blocks where participants watch and listen to blurred videos with degraded, incomprehensible speech; the reading-based comprehension task includes blocks where participants read sentences and control blocks where they read nonwords, with each sentence or list of nonwords followed by a memory probe to ensure alertness. The multiple demand (MD) localizer task includes blocks where participants solve harder (dark blue) vs. easier (light blue) arithmetic addition problems. C/ Functional localization approach in example participants: language and MD parcels or “search spaces” were intersected with each participant’s activation map for the respective localizer contrast and the top 10% of most responsive voxels within each parcel were selected as the fROIs for that individual. Responses to the language and MD conditions were then extracted from these individually defined language and MD fROIs using a left-out portion of the data—not used for fROI definition—to ensure statistical independence (e.g.,^127^). PWA: participants with aphasia. **Alt text**: Overview of the study. A table compares 37 people with aphasia and 38 controls, next to a left-hemisphere lesion overlay. Example trials show three localizer tasks: audio-visual sentences, reading sentences and arithmetic additions. Brain maps show language regions (red) and multiple demand regions (blue) defined in each individual participant.

**Figure 2.**
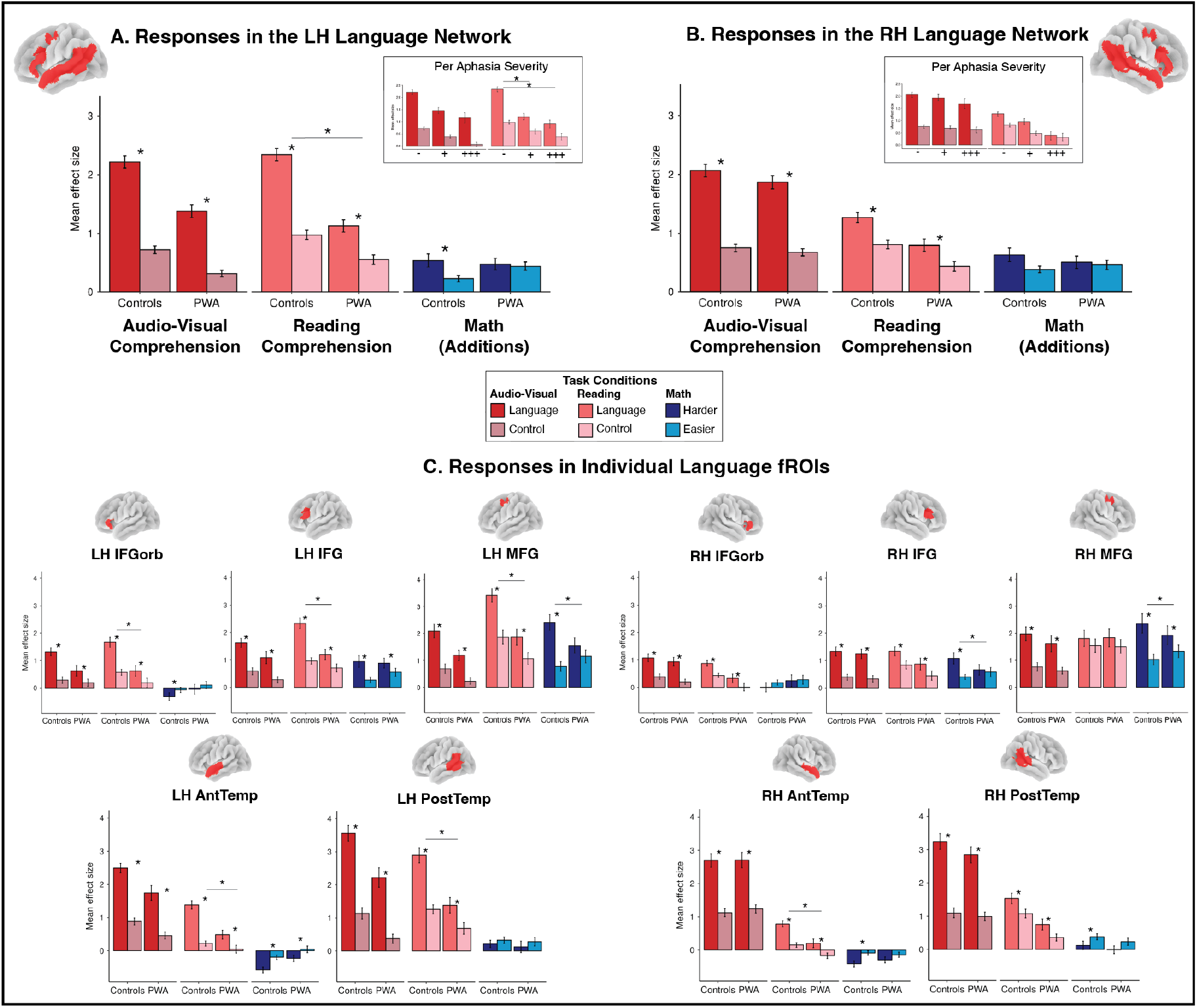
The bilateral language network of participants with aphasia (PWA) responds selectively during language comprehension in both audio-visual and reading modalities. A-B/ Responses of the language network to the audio-visual language comprehension (dark/light red), reading-based language comprehension (dark/light pink), and arithmetic addition task (dark/light blue) contrasts at the hemisphere level (A: LH, B: RH) and the individual fROI level (C) in participants with post-stroke aphasia (PWA) and age-matched controls. Insets in (A-B) show language responses by aphasia severity: controls (-), PWA with recovered to mild aphasia (+, AQ ≥ 76), and PWA with moderate to severe aphasia (+++, AQ < 76). In the left hemisphere, the reading effect was reduced in both PWA subgroups relative to controls, with no difference between PWA subgroups. No subgroup differences were found in the right hemisphere. Results for PWA with WAB-AQ < 93.8 are presented in Supplementary Fig. 1. The language fROIs were defined using the audio-visual language localizer; the results using the reading localizer are highly similar and presented in **Supplementary** Fig. 3. Asterisks represent significant differences (Holm-corrected at the hemisphere level, FDR-corrected at the fROI level) between task conditions (α=0.05), lines with asterisks represent significant differences between groups (α=0.05). **Alt text:** Bar graphs of language network responses. Both groups respond more to sentences than to control conditions. In the left hemisphere these responses are reduced in people with aphasia, especially during reading, regardless of aphasia severity. Right-hemisphere responses are more similar across groups.

### fMRI Experimental Design

Participants completed three tasks while undergoing fMRI (**Fig. 1B**): two language ‘localizer’ paradigms (an audio-visual version and a reading-based version) designed and previously validated to identify the language network within individual participants and measure its response properties (e.g., ^30,97,116,128^); and an MD network ‘localizer’ paradigm (an arithmetic addition task) designed and previously validated to identify and measure response properties of the MD network^85,129,130^. We included two versions of the language localizer for two reasons: to maximize our chances of reliably identifying language areas in each individual, given that different versions may work better for different participants; and to generalize our findings across the listening and reading modalities. We had attempted to include an alternative version of the MD localizer as well (a spatial working memory task; e.g., ^95^), but the task proved too difficult for PWA (with accuracies of ∼60% on both the harder and easier conditions), so we chose not to include the data from this task. All paradigms used a blocked design, and each participant completed two runs of each task, with condition order counterbalanced across runs. The scanning session lasted approximately two hours including breaks. Prior to the scanning session, participants practiced the tasks on a laptop.

In the audio-visual language localizer, participants watched short videos. The videos for the critical condition consisted of a woman producing sentences (all 12 words long and varying in structure and meaning; the sentences were extracted from language corpora and recorded with the help of a trained actress and a professional videographer). For the control condition, the video was blurred and the audio distorted to render speech incomprehensible^124^. In the reading-based language localizer task, participants read eight-word-long sentences (critical condition) and sequences of eight nonwords (control condition), presented one word/nonword at a time ^75,116^. At the end of each trial, a word/nonword was presented, and participants had to decide whether it was the same as the last word/nonword of the trial. The task was designed to be easy and was included to help participants stay alert (see ^108^ for evidence that the language network’s topography and responses are the same for passive reading vs. this, and other, tasks).

In the MD localizer task, participants solved two-operand addition problems that were either harder (critical condition, involving double-digit addends: e.g., 36+45 = 81) or easier (control condition, involving single-digit addends: e.g., 2+4 = 6). The problems were shown with the sum, and the sum was correct on half of the trials. Participants responded with a button press to indicate whether the sum was correct or incorrect. Slightly different versions of the reading-based language localizer task and the MD task were used by PWA and controls to adjust for differences in processing speed and task performance (see **Supplementary Tables 2-5** for details of the procedure and timing; **Supplementary Tables 6-7** for behavioral performance).

In addition, two naturalistic-cognition paradigms were included (and used in the current study for the functional connectivity analyses): a resting-state run (participants were instructed to close their eyes but stay awake and allow their minds to wander) and a ∼five-minute-long story from the Natural Stories corpus (^131^; an edited version of the publicly available story “Elvis Died at the Florida Barber College” by Roger Dean Kiser; unedited version: www.eastoftheweb.com/short-stories/UBooks/ElvDies.html). The story was edited to incorporate various linguistic features known to increase processing difficulty based on behavioral investigations of incremental sentence processing and has been used in numerous prior fMRI studies^95,103–105^. The story was played twice during the scanning session, and the results were averaged across the repetitions in the analyses. The story was followed by seven yes/no content-related questions, included to assess comprehension levels (PWA mean accuracy = 78% ± 22, Controls mean accuracy = 97% ± 11).

### MRI Acquisition

Images were acquired on two identical scanners with identical sequences at Boston University and Massachusetts Institute of Technology (a Siemens 3.0T Prisma-upgraded scanner with a 32-channel head coil). Structural T1-weighted and functional BOLD data were collected for all participants. In particular, anatomical (T1-weighted) data were acquired using a T1 MEMPRAGE sequence (176 sagittal slices, 1mm×1mm×1mm voxel size, TR = 2530ms, TE1 = 1.69ms, TE2 = 3.55ms, TE3 = 5.41ms, TE4 = 7.27ms, TI = 1100ms, flip angle = 7.0 degrees). Functional data were acquired using an SMS EPI sequence (2.4 × 2.4 × 1.8 mm voxels, TR = 2,000 ms, TE = 32 ms, 81 slices). See **Supplementary Methods** for additional details on the sequences.

### fMRI Preprocessing

fMRI data were preprocessed using SPM12 (release 7487), the CONN EvLab module (release 19b), and custom MATLAB scripts. For each PWA, a lesion mask was generated using semi-automated segmentation in ITK-SNAP^132^. Functional scans were coregistered to the first scan of the first run, and potential outlier scans were identified based on motion estimates and BOLD signal indicators using default CONN thresholds. Functional and structural data were normalized to the MNI space (IXI549Space template) using SPM12 unified segmentation with the lesion masked out, resampled to 2 mm isotropic voxels, and smoothed with a 4 mm FWHM Gaussian kernel (excluding voxels within the lesion mask). The data from the naturalistic paradigms underwent additional denoising: nuisance regression of motion parameters and white matter/CSF signals, followed by bandpass filtering (0.01–0.25 Hz).

### First-level Analysis

Voxel-level responses were estimated using a General Linear Model (GLM) in which each experimental condition was modeled with a boxcar regressor convolved with the canonical hemodynamic response function (HRF) (fixation was modeled implicitly, such that all timepoints that did not correspond to one of the conditions were assumed to correspond to a fixation period). Temporal autocorrelations in the BOLD signal timeseries were accounted for by a combination of high-pass filtering with a 128-second cutoff, and whitening using an AR(0.2) model (first-order autoregressive model linearized around the coefficient a=0.2) to approximate the observed covariance of the functional data in the context of Restricted Maximum Likelihood estimation (ReML). In addition to experimental condition effects, the GLM design included first-order temporal derivatives for each condition (included to model variability in the HRF delays), as well as nuisance regressors to control for the effect of slow linear drifts, subject-specific motion parameters (6 parameters), and potential outlier scans (identified during preprocessing as described above) on the BOLD signal.

### Functional Localization of Networks and Response Estimation

For each participant, functional regions of interest (fROIs) were defined using the Group-constrained Subject-Specific (GSS) approach^116,133^, whereby a set of parcels or “search spaces” (i.e., brain areas within which most healthy individuals in prior studies showed activity for the corresponding contrast) was intersected with each participant’s activation map for the respective localizer contrast. To define the language fROIs, we used five parcels derived from a group-level probabilistic activation overlap map for the sentences > nonwords contrast in 220 independent participants^98,125,134–136^. These parcels include three regions in the left frontal cortex: two located in the inferior frontal gyrus (LH IFG and LH IFGorb), and one located in the middle frontal gyrus (LH MFG); and two regions in the left temporal cortex spanning the extent of the lateral temporal lobe (LH AntTemp and LH PostTemp). Additionally, we examined activations in the right hemisphere (RH) homotopes of the language regions. To define the RH fROIs, the LH parcels were mirror-projected onto the right hemisphere to create five homotopic parcels. By design, the parcels cover relatively large swaths of cortex to accommodate inter-individual variability, making their mirrored versions likely to encompass right-hemisphere language regions despite potential hemispheric asymmetries in the precise locations of activations^75,137^. Individual language fROIs were defined by selecting within each parcel the 10% of most localizer-responsive voxels based on the t-values for the intact > degraded speech contrast of the audio-visual language task or the sentences > nonwords contrast of the reading-based task (see **Fig. 1C**). Analyses were performed using each set of language fROIs for each individual (audio-visual and reading-based). To define the MD fROIs, we used a set of 20 parcels (10 in each hemisphere) derived from a group-level probabilistic activation overlap map for the hard > easy spatial working memory contrast in 197 participants, which have been used in numerous prior studies ^85,95,105,107^ *inter alia*. These parcels include regions in the parietal cortex (PostParietal, MidParietal, AntParietal), superior frontal gyrus (SFG), precentral gyrus (PCG), IFG pars opercularis (IFGop), middle frontal gyrus and its orbital part (MFG, MFGorb), insula, and medial frontal cortex (Medial Front). Individual MD fROIs were defined by selecting within each parcel the top 10% of most localizer-responsive voxels based on the t-values for the hard > easy contrast of the MD localizer task.

The language and MD parcels are available at https://www.evlab.mit.edu/resources-all/download-parcels. In PWA, an fROI was excluded when ≥ 90% of the corresponding parcel overlapped with the participant’s lesion mask. Overlap was calculated as the intersection between the lesion mask and the parcel (see **Supplementary Table 8** for the number of PWA included per fROI and task).

For both the language and the MD networks, we estimated the responses in the individually defined fROIs to the intact and degraded speech conditions of the audio-visual language task, to the sentences and nonwords conditions of the reading task, and to the easy and hard conditions of the arithmetic addition task. For extracting the responses of the language fROIs to the same language task conditions, and for extracting the responses of the MD fROIs to the arithmetic task conditions, an across-runs cross-validation procedure was used (first, data from run 1 were used to define the fROIs and data from run 2—to estimate the responses; second, data from run 2 were used to define the fROIs and from run 1—to estimate the responses; finally, the response estimates were averaged to obtain a single value per condition per fROI per participant). This approach ensures statistical independence between the data used for fROI definition and response estimation^127,138^. Statistical tests were performed on these estimate values.

### Estimation of Inter-regional Functional Correlations during Naturalistic Cognition Paradigms

Analyses of inter-regional functional correlations, typically referred to as functional connectivity (FC), were performed on the naturalistic paradigms data (i.e., resting state and story listening). First, for each naturalistic run, the average BOLD signal intensity time-course was extracted from each language and MD fROI (defined as explained above). Then, for each pair of language fROIs, MD fROIs, and language-MD fROIs, the Fisher-transformed Pearson’s correlation coefficient was computed between their respective time-courses using custom R scripts (R version 4.2.1). For participants with two story-listening runs, correlation coefficients were averaged across runs. These analyses resulted for each naturalistic paradigm and for each participant in a 10×10 correlation matrix of functional connectivity among the language regions, a 20×20 correlation matrix of functional connectivity among the MD regions, and a 10×20 correlation matrix of functional connectivity between the language and MD regions. Statistical tests were performed on these correlation values.

### Behavioral Assessments

A battery of standardized assessments was administered to PWA to assess their language and cognitive abilities. These assessments included the Western Aphasia Battery-Revised (WAB-R)^126^ to characterize aphasia severity, the Boston Naming Test (BNT) to assess naming abilities^139^, the Psycholinguistic Assessments of Language Processing in Aphasia (PALPA) to assess lexical decision and non-word repetition^140^, the three-picture version of the Pyramids and Palm Trees Test (PAPT) to evaluate non-verbal semantic abilities^141^, and the Reading Comprehension Battery for Aphasia - second edition (RCBA)^142^ to assess general reading abilities at the sentence and word levels. Auditory and visual sentence comprehension were further evaluated using a computerized version of a sentence-picture matching task developed by Zimmerer and colleagues^143^. In addition, general cognitive skills including attention, memory, and visuospatial processing were evaluated with the Repeatable Battery for the Assessment of Neuropsychological Status (RBANS)^144^, the Wechsler Memory Scale fourth edition (WMS-4)^145^, and the Cognitive Linguistic Quick Test (CLQT)^146^. All assessments were administered over videoconference or in person depending on the participant’s ability to use technology and COVID-19 pandemic safety protocols.

### Statistical Analyses

We investigated group differences between PWA and controls across two primary neural measures: (1) responses within the language and MD fROIs during the two language tasks (audio-visual and reading-based) and the arithmetic addition task; and (2) functional connectivity (FC), defined as the Pearson’s correlation of BOLD time-courses between fROI pairs during the two naturalistic paradigms (resting state and story comprehension).

#### Task activation analyses

Overall, group comparisons were conducted using linear mixed-effects models (LMEMs) in R fitted with the bobyqa optimizer in *lme4*^147^, with Satterthwaite degrees of freedom via *lmerTest*^148^ (also used for *emmeans* contrasts). The random-effects structure followed the design of the data^149^: random intercepts for participants and fROIs, by-participant random slopes for the within-participant effect being tested (response condition or task), and a random intercept for each participant’s fROI, since the conditions were measured within the same fROI. By-fROI random slopes were not included, as inference was made over participants for a predefined set of fROIs. When a model was singular or did not converge, correlations between random effects were first removed and, if needed, variance components estimated at zero were dropped; the by-participant slopes of the tested effects were always retained. Confidence intervals were computed by the Wald method, and marginal/conditional R^2^ via the *performance* package.

Within each group, the language > control effects (intact > degraded speech for the audio-visual task, sentences > nonwords for the reading task) and the hard > easy arithmetic effect were first tested separately for each LH and RH language and MD fROI using paired t-tests when a Shapiro-Wilk test indicated normality of the pairwise differences (p ≥ 0.05) and Wilcoxon signed-rank tests otherwise (effect size: Cohen’s d or r = Z/√N, respectively), FDR-corrected across the fROIs tested together within each localizer × task × group combination. At the hemisphere level, each contrast was tested within each group with an LMEM (Effect size ∼ Response condition + (1 + Response condition | participant) + (1 | fROI) + (1 | participant:fROI)), fitted separately for the LH and RH fROIs of each localizer × task combination, and Holm-corrected across all localizer × task × group × hemisphere combinations.

Group differences in each contrast were tested at the hemisphere level with an interaction LMEM (Effect size ∼ Response condition * Group + (1 + Response condition | participant) + (1 | fROI) + (1 | participant:fROI)), Holm-corrected across all localizer × task × hemisphere combinations, and at the level of individual fROIs with a simpler model (Effect size ∼ Response condition * Group + (1 | participant)), in which the participant intercept accounts for the pairing of the two conditions within the fROI, FDR-corrected across fROIs within each localizer × task combination.

Functional selectivity (each language contrast vs. the hard > easy arithmetic contrast in the language fROIs, and the reverse in the MD fROIs) was tested using per-participant, per-fROI task contrasts. At the hemisphere level, these were modeled within each group with an LMEM (Contrast ∼ Task + (1 + Task | participant) + (1 | fROI) + (1 | participant:fROI)) and *emmeans* contrasts, FDR-corrected across all localizer × group × hemisphere combinations. At the fROI level, the contrasts were compared with the paired t-test/Wilcoxon procedure described above, FDR-corrected across fROIs within each localizer × group combination. To test whether responses differed by aphasia severity, patients were stratified using the Aphasia Quotient (AQ) from the Western Aphasia Battery (AQ ≥ 76 = mild aphasia; AQ < 76 = moderate to severe aphasia), and the hemisphere-level interaction model was refit for each localizer × task × hemisphere combination with this three-level group factor (controls, mild, moderate-severe) in place of the binary group factor. Pairwise group differences (interaction contrasts, and simple contrasts within the preferred condition) were extracted via *emmeans* and Holm-corrected separately within the interaction contrasts and within the simple contrasts.

The analyses of the response profiles were complemented with analyses of spatial overlap between the language and the MD network, and analyses of lateralization for the language network (see **Supplementary Material**).

#### Functional connectivity analyses

Fisher z-transformed Pearson correlations between fROI-pair BOLD time-courses were modeled with an LMEM including random intercepts for participant and fROI pair. To test for network segregation (i.e., whether within-network connectivity was stronger than between-network connectivity), an LMEM with network type (within-language-network, within-MD-network, between-network) as fixed effect, by-participant random slopes for network type (following the simplification procedure described above), and random intercepts for participant and fROI pair was fit separately for each group, task, and hemisphere (within-LH, within-RH, interhemispheric); pairwise contrasts were extracted via *emmeans* with Tukey correction. Group comparisons were performed separately for each naturalistic paradigm (resting state and story) and each of the network × hemisphere combinations using an LMEM (Corr ∼ Group + (1|participant) + (1|fROIpair)); p-values were FDR-corrected across all tests.

#### Brain-Behavior Analyses

For this component of the study, we pre-registered a set of planned analyses to evaluate the relationship between a) neural measures extracted from the language and MD networks and b) behavioral measures of linguistic and cognitive abilities in PWA (https://osf.io/xcfpz/). Prior to all analyses, variables were z-scored, and missing data were imputed using predictive mean matching (MICE, 50 iterations). To reduce the dimensionality of the behavioral dataset while retaining meaningful variance, we first carried out a Principal Component Analysis (PCA) on the behavioral assessments described above (see **Supplementary Methods**); the resulting components served as the dependent variables for the analyses described below.

For the neural data, we used two complementary approaches:

1. The *PCA approach*: We applied a PCA to reduce the dimensionality of neural features across participants (see **Supplementary Methods**). We then used multiple linear regression to test relationships between these neural components and the behavioral principal components (i.e., BC_i_ ∼ NC1 + NC2 + … + NC_K_ + lesion volume, where BC_i_ represents the i^th^ behavioral component and K is the number of neural components NC).
2. The *Elastic Net approach*: To provide more granular insights into specific neural predictors, we employed a two-stage approach. Variable selection was performed using elastic net regression with leave-one-out cross-validation, separately for three pre-defined neural feature sets: language network features, MD network features, and language-to-MD functional connectivity features, with lesion volume as a candidate predictor in each set. The mixing parameter was fixed at α = 0.5 (equal L1 and L2 penalization), and λ was the value minimizing the cross-validated mean squared error. Predictors with non-zero coefficients at this λ were then entered into confirmatory linear regression models to obtain inferential statistics. Because variable selection and confirmatory regression were performed on the same data, the confirmatory R² and p-values may be optimistic; we therefore also report the leave-one-out cross-validated R² of the elastic net (1 − cross-validated mean squared error / outcome variance).

For both approaches, residual normality (Shapiro-Wilk) and homoscedasticity (Breusch-Pagan) were tested and multicollinearity was checked (variance inflation factors); when either test was significant (p < 0.05), heteroscedasticity-consistent (HC1) robust standard errors were used, with a robust Wald F test for the overall model. FDR correction was applied to the two overall model p-values in the PCA-based analyses, and across all predictors of the confirmatory regression models.

The relationship between aphasia severity and engagement of the MD and language networks during language processing was assessed with four LMEMs (two networks [language, MD] x two language contrasts [intact > degraded speech, sentences > nonwords]), with fROI-level effect sizes as the outcome, WAB-AQ as the predictor, and participants and fROIs as random intercepts. Because model residuals were heavy-tailed, all models were fitted as robust LMEMs (*robustlmm* package), with p-values based on a normal approximation; standard LMEMs (*lmerTest*, Satterthwaite degrees of freedom) yielded the same conclusions. A secondary, exploratory analysis used the same approach for the response to each language condition (intact speech, sentence reading) relative to fixation. FDR correction was applied within each family of four tests (primary contrasts, fixation responses, hemisphere follow-ups). All analyses were conducted in R using the *lme4*, *lmerTest*, and *robustlmm* packages.

Given that previous work has found distinct patterns of reorganization and recovery following frontal versus temporal lesions ^150–154^, we examined whether lesion *location* modulated the relationship between neural measures and linguistic abilities. We fitted linear regression models testing two-way interactions between each neural component (NC from the PCA) and lesion location (i.e., proportion of damage in frontal / temporal language parcels) in predicting the behavioral component (BC) reflecting linguistic abilities. The models took the form: BC1 ∼ NC_j_*(Frontal Damage + Temporal Damage), where NC_j_ represents each neural component, Frontal Damage represents the mean proportion of damage across LH IFGorb, IFG and MFG language parcels, and Temporal Damage represents the mean proportion of damage across LH anterior and posterior temporal language parcels. Influence diagnostics applied to each model separately identified outliers based on Cook’s distance > 4/n; models were refit without these cases. To assess robustness, we report results both with and without these outlier cases.

In a final exploratory analysis, we tested whether the relationship between responses to language in the MD network and linguistic abilities depended on lesion location, with models of the same form using the intact > degraded speech and sentences > nonwords effects averaged across the MD fROIs (excluding fROIs with ≥ 90% damage): BC1 ∼ Mean Language > Control Effect × (Frontal Damage + Temporal Damage), with the same outlier procedure. As a follow-up, the same models were fitted with the MD network responses to sentence reading and to nonword reading relative to fixation.

## Results

### The response profile of the language network

The language network of PWA showed a strong and selective response to language comprehension in both audio-visual and reading modalities, but the response was weaker than in age-matched controls in the left hemisphere (**Fig. 2**). Here, we report the results for the language fROIs defined using the audio-visual language localizer, but the results are highly similar when the fROIs are defined with the reading-based localizer (**Supplementary Fig. 3**).

All left-hemisphere (LH) language fROIs responded more strongly during the language comprehension condition than the control condition in both PWA and age-matched controls in both modalities (FDR-controlled for the number of fROIs; all *p_fdr_* < 0.008, **Fig. 2A,C**). Similarly, in both groups, all homotopic right-hemisphere (RH) language fROIs showed a language > control effect in both modalities (all *p_fdr_* < 0.04, **Fig. 2B,C**), with the exception of the RH MFG fROI’s response to the reading-based contrast in control participants (*p_fdr_* = 0.105).

In the left hemisphere, at the network level, PWA showed an overall weaker response than controls to the reading contrast (t(74) = −5.08, *p_holm_* < 0.001, d = −0.57); the response to the audio-visual contrast was also weaker, but this difference did not survive correction (t(69) = −2.92, p = 0.005, *p_holm_* = 0.061, d = −0.32; **Fig. 2A**). At the fROI level, PWA showed a weaker language > control effect for the reading contrast in all five LH fROIs (all *p_fdr_* < 0.012, **Fig. 2C**). This was not the case for the audio-visual contrast, for which responses were lower in magnitude in most left fROIs but the language>control effect did not statistically differ from controls (all *p_fdr_* > 0.06).

In the right hemisphere, at the network level, there were no significant group differences in language > control effect in either modality (audio-visual: t(72) = −0.76, *p_holm_* = 1.00; reading: t(73) = −0.75, *p_holm_* = 1.00; **Fig. 2B**). However, at the fROI level, PWA showed a weaker language > control effect in the reading modality in the AntTemp fROI only (*p_fdr_*=0.034, see **Fig. 2C**), with no reliable group differences observed in the audio-visual modality (all *p_fdr_* > 0.30). For detailed results of hemisphere-level group comparisons in language responses across modalities and localizers, see **Supplementary Table 9**.

Consistent with these hemisphere-specific differences, the language network of PWA was less left-lateralized than that of controls during the audio-visual (PWA: LI = −0.13 ± 0.46; controls: LI = 0.11 ± 0.21; *p_fdr_* = 0.006), but not during the reading task (PWA: LI = 0.39 ± 0.58; controls: LI = 0.51 ± 0.34; *p_fdr_* = 0.90; **Supplementary Fig. 4**). This change reflected reduced left-hemisphere engagement rather than increased right-hemisphere recruitment: during the audio-visual task, PWA had fewer suprathreshold voxels than controls in the left-hemisphere language parcels and not more, if anything fewer, in the right-hemisphere parcels. During the reading task, suprathreshold voxel counts were reduced in both hemispheres, leaving the lateralization index unchanged.

In both groups, language network responses were strongly selective for language in both hemispheres, with significantly larger language contrasts (in both modalities) than MD contrasts (hard > easy arithmetic) (all *p_fdr_* < 0.05). In particular, in PWA the MD contrast was non-significant in the language network of each hemisphere (all *p_holm_* = 1.00). At the level of individual fROIs, a weak positive hard > easy effect was found in two frontal fROIs (LH IFG: *p_fdr_* = 0.023; RH MFG: *p_fdr_* = 0.021). However, this effect was not specific to PWA: controls showed a positive hard > easy effect in the same frontal fROIs, as well as in the LH MFG and RH IFG fROIs (all *p_fdr_* < 0.001). The effect was never significantly larger in PWA than in controls, and it was reduced in PWA in the LH MFG (*p_fdr_* < 0.001), RH IFG and RH MFG (both *p_fdr_* = 0.037) fROIs. Thus, the response of the language network to the arithmetic task did not differ qualitatively between groups, and there was no evidence of increased recruitment of the language fROIs for non-linguistic processing in PWA.

### The response profile of the Multiple Demand network

The bilateral MD network of PWA did not show any language-selective responses, similar to controls (**Fig. 3**). In particular, both PWA and controls showed close to zero or negative responses to the audio-visual language conditions (intact and degraded speech; no intact > degraded effect at the hemisphere level in either group, all *p_holm_* = 1.00) and low responses to the reading conditions. The response to nonword reading was numerically stronger than to sentence reading in both groups and hemispheres—the opposite of the language network’s profile—but this effect was significant only in the right hemisphere of PWA (*p_holm_* = 0.029; controls LH: *p_holm_* = 0.12, RH: *p_holm_* = 0.075; PWA LH: *p_holm_* = 0.64). It did not differ significantly between groups in either hemisphere (both *p_holm_* = 1.00) (**Fig. 3A-B**). Even when restricting the analyses to the subgroup of individuals with more severe aphasia (i.e., WAB-AQ < 76), responses to the language conditions in the MD fROIs did not differ significantly from those of controls (all *p_holm_* = 1.00, see insets in **Fig. 3A-B** and **Supplementary Table 10**). In contrast, the MD network across both hemispheres, and all individual MD fROIs, showed a robust response to the hard > easy arithmetic addition contrast in both groups (all *p_holm/fdr_* < 0.001, **Fig. 3C**). However, similar to the language network’s response to the language tasks, PWA showed an overall weaker response to the arithmetic task contrast than the controls in both hemispheres (LH: t(73) = −4.85, *p_holm_* < 0.001, d = −0.66; RH: t(73) = −3.83, *p_holm_* = 0.004, d = −0.53, **Fig. 3A-B**); this effect held in the individual fROIs (all *p_fdr_* < 0.02, **Fig. 3C**), except for the RH MFGorb fROI (*p_fdr_* = 0.063).

**Figure 3.**
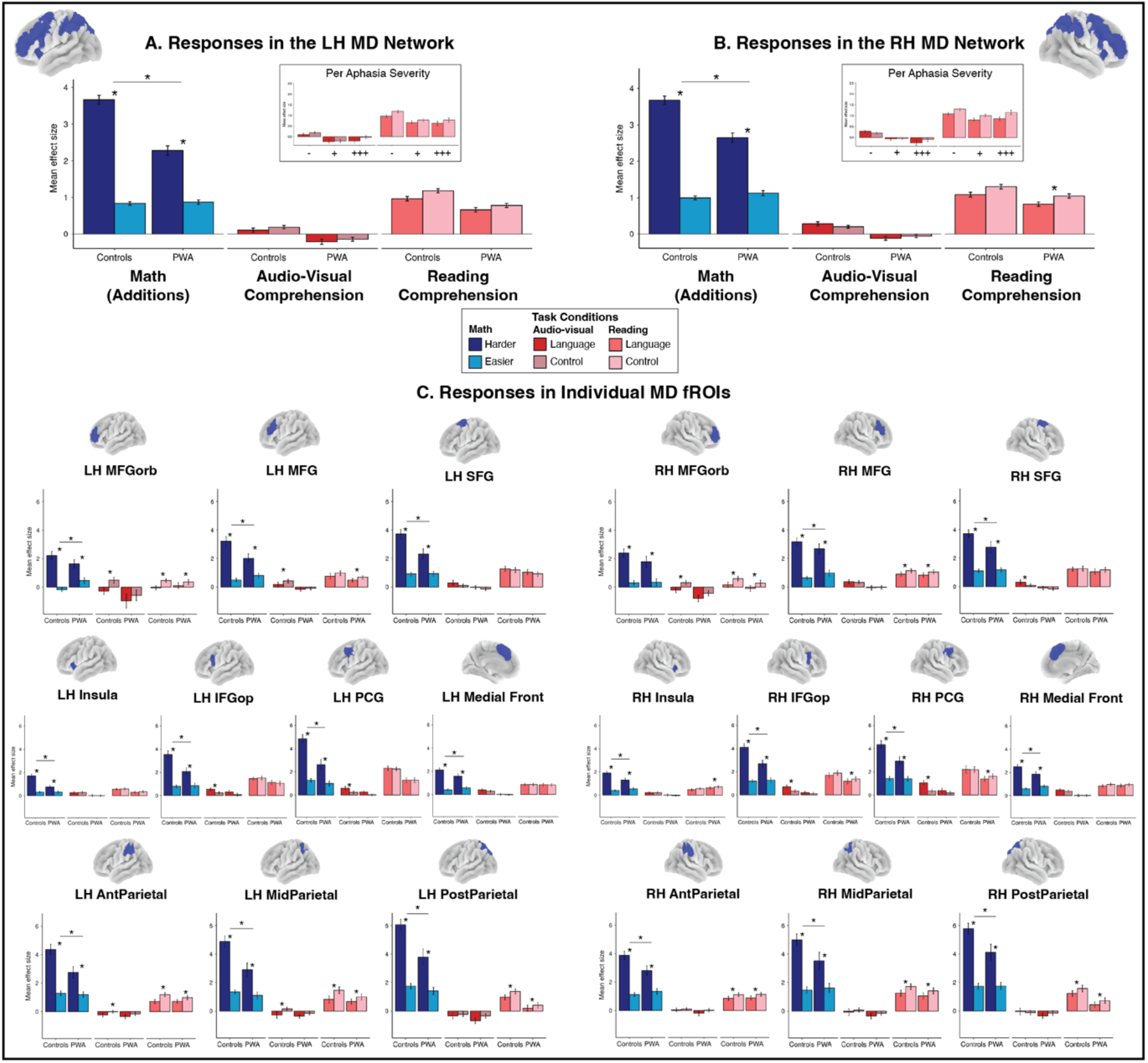
The MD network does not support language comprehension in individuals with chronic post-stroke aphasia, similar to controls. A-B/ Responses of the MD network to the MD (Hard and Easy, dark/light blue), audio-visual language comprehension (dark/light red), and reading-based language comprehension (dark/light pink) conditions at the hemisphere (A: left, B: right) and the individual fROI (C) levels in participants with post-stroke aphasia (PWA) and age-matched controls. Insets in (A-B) show no significant differences between aphasia severity groups in the responses to language contrasts in the MD fROIs of each hemisphere: controls (-), PWA with recovered to mild aphasia (+, AQ ≥ 76), and PWA with moderate to severe aphasia (+++, AQ < 76). Asterisks represent significant differences (Holm-corrected at the hemisphere level, FDR-corrected at the fROI level) in effect size between conditions (α=0.05), lines with asterisks represent significant differences in contrast magnitude between groups (α=0.05). **Alt text:** Bar graphs of multiple demand network responses. In both hemispheres, both groups respond more to the harder than to the easier math condition, but with overall reduced responses in people with aphasia. Responses to language tasks are near zero or larger for the control condition in both groups and at every aphasia severity level. The same pattern appears in nearly all 20 individual regions.

In the controls, several MD fROIs (bilateral IFGop and PCG fROIs, and the RH SFG fROI) showed a significant audio-visual language effect (all *p_fdr_* < 0.04, **Fig. 3C**). However, these effects a) did not generalize to the reading modality, which argues against their interpretation as reflecting language processing; and b) were small in size and significantly smaller in magnitude than the hard > easy arithmetic effect in the same fROIs (all *p_fdr_* < 0.001).

Critically, these language>control effects were not observed in PWA: no MD fROI showed a significant intact > degraded speech or sentences > nonwords effect.

### The language-MD network dissociation in PWA in patterns of response, spatial overlap, and functional connectivity

The dissociation between the language and MD networks in PWA is evident in three complementary measures: task activation patterns in individually defined fROIs, spatial overlap of thresholded activation maps, and inter-regional functional connectivity during naturalistic paradigms. First, language and MD fROIs showed distinct profiles even for adjacent regions within the left frontal cortex (**Fig. 4A**). The left frontal language fROIs showed robust responses to both language comprehension contrasts (all *p_fdr_* < 0.008), whereas the neighboring MD fROIs showed the opposite profile: strong hard > easy arithmetic effects (all *p_fdr_* < 0.001) and no language > control effect in either modality. The left IFG and MFG language fROIs also responded to the arithmetic conditions, and the hard > easy contrast reached significance in the IFG fROI only (*p_fdr_* = 0.023; MFG: *p_fdr_* = 0.061; IFGorb: *p_fdr_* = 0.80; **Fig. 2C**). In the IFG fROI, the audio-visual language effect was larger than the arithmetic effect (*p_fdr_* = 0.043), but the reading effect was not (*p_fdr_* = 0.75).

**Figure 4.**
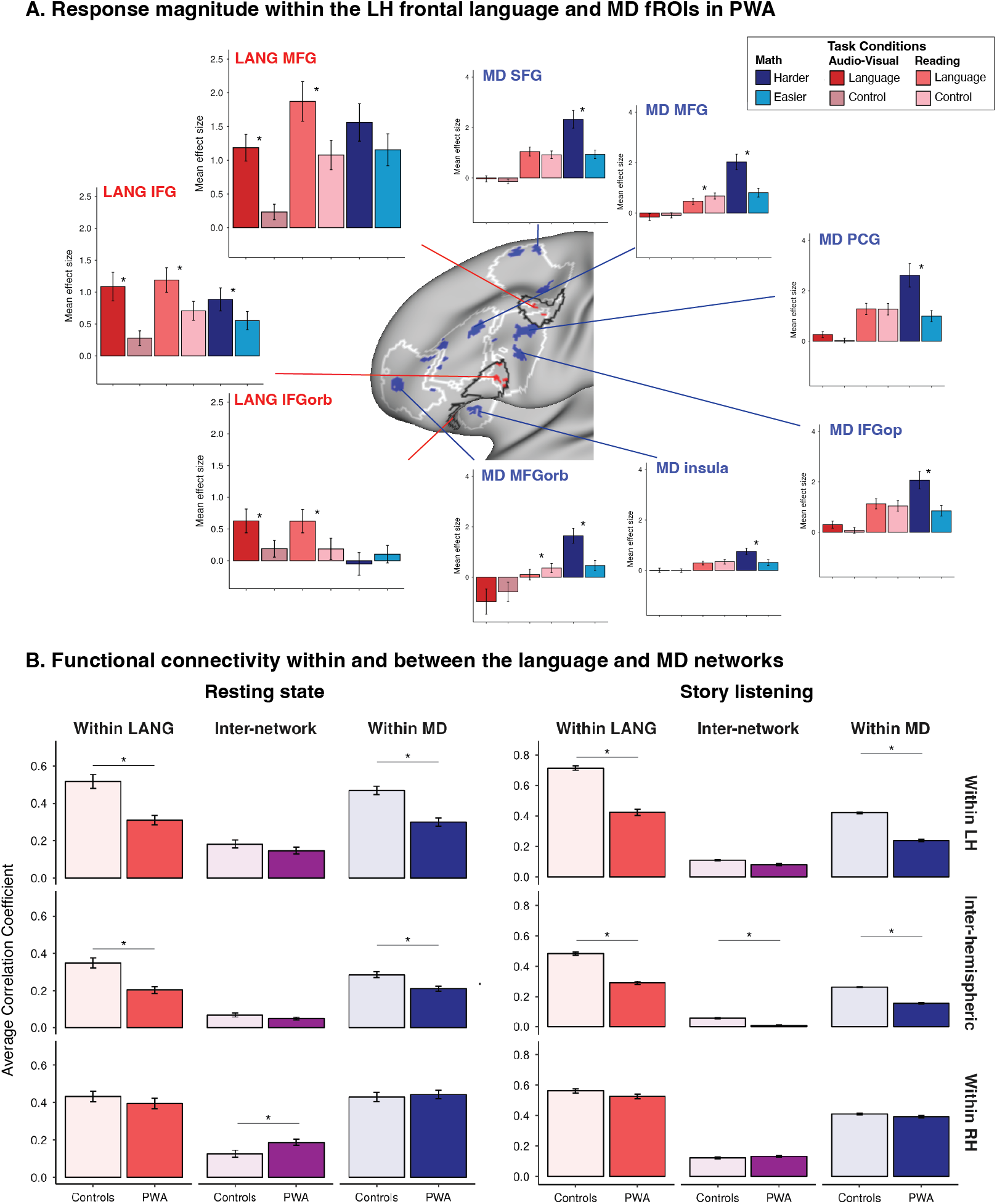
The language and the MD networks remain robustly dissociated in PWA, similar to controls. A/ Response magnitude to the reading-based language comprehension (sentences > nonwords), audio-visual language comprehension (intact speech > degraded speech), and MD math (harder > easier arithmetic additions) task conditions in the left hemisphere (LH) language (left plots) and MD (right plots) frontal fROIs in participants with post-stroke aphasia (PWA). This plot is showing a subset of the data in Figs. 2 and 3 for easier comparison. The LH language frontal fROIs of PWA responded robustly to both language contrasts, with a weak hard > easy contrast effect in the IFG fROI only; while the LH MD frontal functional ROIs (fROIs) showed a strong task-difficulty effect but no significant response to either language contrast. White outlines on the cortical surface represent MD frontal parcels and black outlines represent language frontal parcels. Dark blue regions represent example MD fROIs (top 10% voxels most responsive to the MD task contrast) and red regions represent example language fROIs (top 10% voxels most responsive to the audio-visual language task contrast) for one representative participant. Asterisks represent significant differences in effect size between conditions (α=0.05, FDR-corrected). B/ Inter-regional correlations in BOLD timeseries between individual-specific language and MD fROIs during naturalistic (rest and story listening) fMRI tasks at the hemisphere level. Lines with asterisks represent significant differences between groups (α=0.05). All significance levels were measured after FDR correction. RH: right hemisphere. **Alt text**: Top bar graphs show that in people with aphasia, left frontal language regions respond preferentially to language and multiple demand regions respond preferentially to harder math, with little overlap. Bottom bar graphs show that connectivity within both networks is lower in people with aphasia within the left hemisphere and across hemispheres across naturalistic paradigms. Right-hemisphere connectivity is largely preserved in people with aphasia and increased between networks during rest.

Second, if the MD regions take on some language function in chronic aphasia, the two networks should show an increase in spatial overlap in PWA. This is not what we observed (**Supplementary Fig. 5**). The Dice coefficient between the thresholded language and MD activation maps was lower in PWA than in controls across all bilateral parcels (audio-visual: *p_fdr_* = 0.003; reading: *p_fdr_* = 0.008) and within the frontal parcels (audio-visual: *p_fdr_* < 0.001), and did not differ between groups for the reading contrast maps within the frontal parcels (*p_fdr_* = 0.051).

And third, functional connectivity analyses aligned with the activation-based analyses in showing clear segregation between the two networks in PWA, similar to controls (**Fig. 4B**). Within-network synchronization was consistently stronger than between-network synchronization for both the language and MD networks during both rest and story listening (for within-left-hemisphere, within-right-hemisphere, and interhemispheric connections: all Tukey-adjusted *p* < 0.02). Relative to controls, PWA showed weakened within-network synchronization within the left hemisphere and between hemispheres across both paradigms (**Fig. 4B**, all *p_fdr_* < 0.05). Critically, however, synchronization between the language and the MD networks did not differ between the groups, or was weaker in PWA, in the left hemisphere and between hemispheres, at rest and during story comprehension (interhemispheric, story: *p_fdr_* = 0.046; all others *p_fdr_* > 0.20). In the right hemisphere, PWA showed a small increase in between-network synchronization at rest (*p_fdr_* = 0.047), but this effect did not generalize to the story comprehension condition (*p_fdr_* = 0.71, **Fig. 4B** left panel).

### Brain-behavior associations

A principal component analysis (PCA) on the standardized assessments in PWA revealed two behavioral components (BC) (see **Supplementary Table 11** for the list of test scores included in the PCA and **Supplementary Methods** for more details on the PCA). BC1 showed strong loadings from linguistic measures, as well as from verbally mediated memory and attention measures. BC2 showed strong loadings from nonverbal reasoning, visuospatial processing, visual and working memory, delayed memory and nonverbal semantic processing (**Fig. 5A**). Thus, we took BC1 to reflect linguistic abilities and BC2 general cognitive and executive function abilities. Together, the two components explained 64% of the total variance in the behavioral data (BC1: 39.5%; BC2: 24.8%).

**Figure 5.**
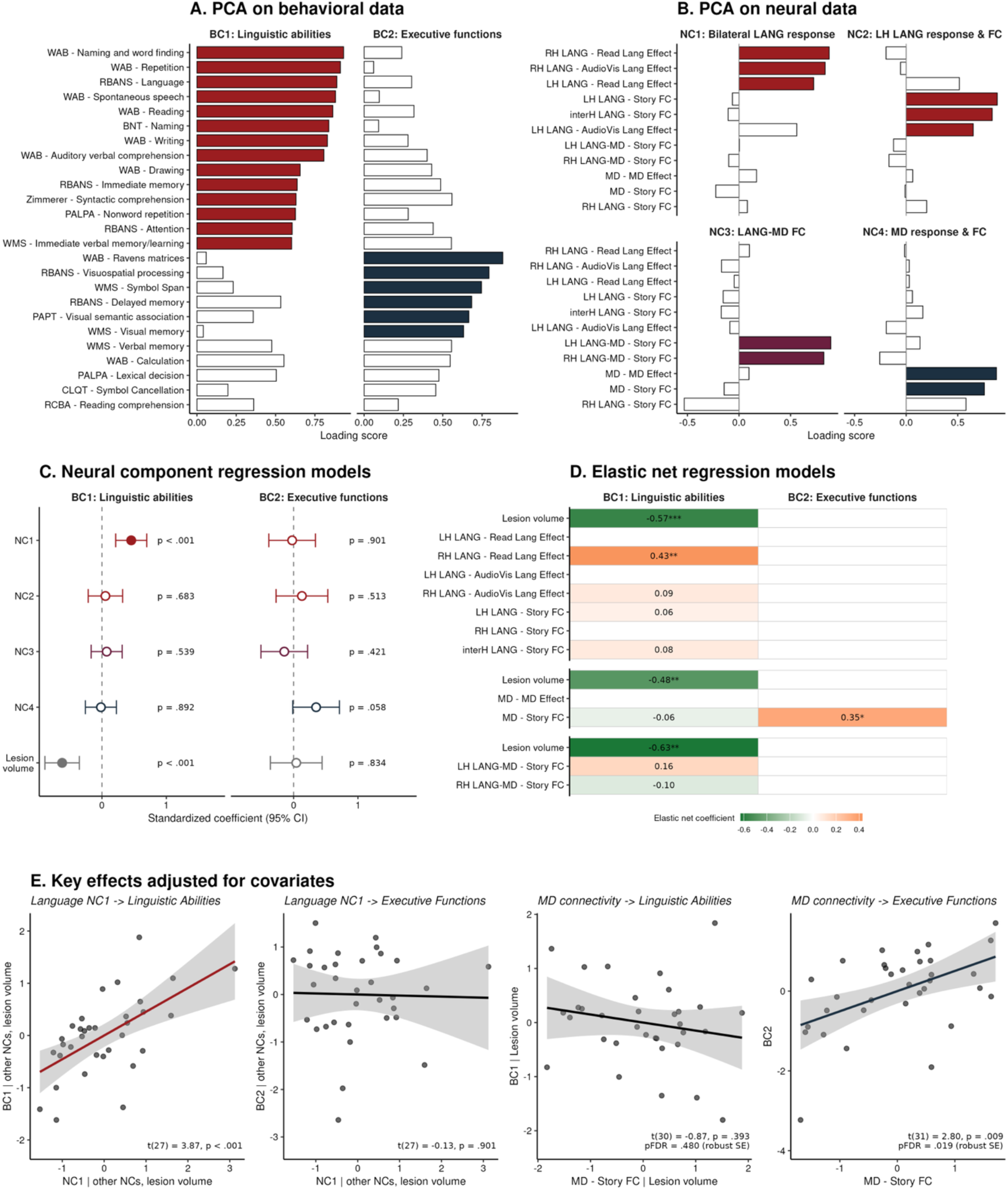
Linguistic abilities in chronic aphasia are best predicted by response strength in the language network, not the MD network. A/ Principal Component Analysis (PCA) loadings derived from behavioral measures (N = 41 PWA, Varimax rotation). Filled bars indicate loadings ≥ 0.60 on the linguistic (BC1) or executive function (BC2) component. B/ PCA loadings derived from neural measures (N = 33 PWA, Varimax rotation; components numbered by explained variance). Filled bars indicate loadings ≥ 0.60. C/ Standardized coefficients (± 95% CI) from the linear regression models predicting each behavioral component from the four neural components and lesion volume. Filled symbols indicate p < 0.05. D/ Elastic net regression models predicting each behavioral component from each neural feature set (language network, MD network, and LANG-MD connectivity features; lesion volume included in each set). Cells show the coefficients of the features selected by the elastic net; blank cells indicate features that were not selected. Asterisks indicate the FDR-corrected significance of the selected features in the confirmatory linear regressions (* *p_fdr_* < 0.05, ** *p_fdr_* < 0.01, *** *p_fdr_* < 0.001). E/ Partial regression plots of the key effects, adjusted for the other predictors of each model: NC1 from the neural component models in C (adjusted for the other neural components and lesion volume), and MD network story-listening connectivity from the elastic net confirmatory models in D (adjusted for the other selected predictors). Lines show the linear fit (± 95% CI); colored lines indicate significant effects. Robust SE: heteroscedasticity-consistent standard errors, applied when model assumptions were violated. AudioVis: audio-visual language task, BC: behavioral component, CI: confidence interval, FC: functional connectivity, FDR: false discovery rate, interH: interhemispheric, LANG: language fROIs, LH: left hemisphere, MD: multiple demand fROIs, NC: neural component, PWA: people with aphasia, Read: reading-based language task, RH: right hemisphere. **Alt text:** Principal component loadings separate behavioral measures into a linguistic and a nonverbal cognitive component. They separate neural measures into four components: two loadings on language network measures (bilateral language response, and left-hemisphere language response and connectivity), one on connectivity between the language and MD networks, and one on MD network response and connectivity. Regression coefficients show that only the bilateral language network component predicts linguistic abilities beyond lesion volume, while no component significantly predicts executive abilities. Elastic net models show that lesion volume and right-hemisphere reading-based language activation predict linguistic abilities, and that MD network connectivity is the only selected predictor of executive abilities. Scatterplots show that the bilateral language network component predicts linguistic but not executive abilities, while MD network connectivity predicts executive but not linguistic abilities.

A PCA on the neural measures (task responses and functional connectivity) in PWA revealed four neural components (NC) (**Fig. 5B**; see **Supplementary Methods** for the list of measures included in the PCA). NC1 and NC2 showed strong loadings from the language network measures. High NC1 scores were associated with stronger language responses in the bilateral language network (sentences > nonwords in the LH and RH, and intact > degraded speech in the RH language fROIs), suggesting that this component reflects the response of the bilateral language network to language. High NC2 scores were associated with stronger functional connectivity within the LH language network and between the LH and RH language networks, together with stronger LH language responses during the audio-visual task, suggesting that this component represents the left-lateralized language network’s integrity. Higher NC3 scores were associated with higher functional connectivity between the language and the MD networks during story listening, suggesting that it reflects the degree of interaction between the two networks. Finally, NC4 showed strong loadings from the MD network measures, such that high NC4 scores were associated with higher MD task responses and functional connectivity in the MD network, along with stronger functional connectivity in the RH language network (loading = 0.58), suggesting that this component reflects properties of the MD network and/or networks not affected by the lesion. Together, the four components explained 76.5% of the total variance in the neural data (NC1: 21.9%; NC2: 20.8%; NC3: 17.1%; NC4: 16.6%).

The two pre-registered brain-behavior analytic approaches revealed convergent evidence for distinct neural features underlying linguistic abilities versus executive functions in PWA (**Fig. 5C-E**). First, linguistic abilities (BC1) were significantly predicted by the model including the four neural components (F(5, 27) = 9.07, *p* < 0.001, *p_fdr_* < 0.001, adjusted R² = 0.56). Among the neural components, only bilateral language network responses (NC1) showed a significant association with linguistic abilities after accounting for lesion volume (β = 0.45, 95% CI [0.21, 0.70], t(27) = 3.87, *p* < 0.001; lesion volume: β = −0.62, t(27) = −4.74, *p* < 0.001). Executive functions (BC2) showed a different pattern: the overall model was not significant (F(5, 27) = 1.01, *p* = 0.43), and the MD network component (NC4) showed a trend-level association with executive functions (β = 0.35, 95% CI [−0.01, 0.71], t(27) = 1.98, *p* = 0.058). The elastic net approach provided convergent evidence with greater feature specificity (**Fig. 5D**; **Supplementary Table 12**). For linguistic abilities, the language network feature set (cross-validated R² = 0.56) had two significant predictors among the selected features: the sentences > nonwords effect in the RH language network (β = 0.51, t(27) = 4.07, *p_fdr_* = 0.002) and lesion volume (β = −0.64, t(27) = −5.64, *p_fdr_* < 0.001). For executive functions, only the MD feature set selected a predictor, MD network connectivity during story comprehension (β = 0.51, t(31) = 2.80, robust SE, *p_fdr_* = 0.019; cross-validated R² = 0.13); no language or language-MD feature was selected.

We also examined the relationship between aphasia severity (WAB-R AQ) and engagement of the language and MD networks during language processing. Aphasia severity was significantly associated only with responses in the language network during the reading comprehension task (sentences > nonwords: β = 0.009 per AQ point, 95% CI [0.005, 0.013], z = 4.21, *p_fdr_* < 0.001), both in the LH (z = 4.04, *p_fdr_* < 0.001) and RH (z= 2.85, *p_fdr_* = 0.009) language fROIs. AQ was not associated with responses during the audio-visual language comprehension task (intact > degraded speech: z = 1.53, *p_fdr_* = 0.25), nor with MD network responses to language in either modality (both *p_fdr_* = 0.98). No significant relationship with AQ survived correction when looking at the magnitude of response to each language condition (intact speech and sentences reading) relative to fixation in either network (all *p_fdr_* > 0.09; **Supplementary Table 13**). Standard LMEMs yielded the same pattern (sentences > nonwords: t(34.3) = 3.33, *p_fdr_* = 0.008; LH and RH follow-ups: both *p_fdr_* = 0.020; all other networks and contrasts *p_fdr_* > 0.10).

### Exploratory analyses of the effect of lesion location on brain-behavior relationships

We next tested whether lesion location (i.e., proportion of damage in frontal vs. temporal language regions) influenced: (1) the relationship between each neural component (NC1–NC4) and linguistic abilities (BC1), and (2) the relationship between language responses in the MD network and linguistic abilities (BC1). The results are reported at uncorrected thresholds given the exploratory nature of these analyses. Across all models, temporal damage consistently predicted poorer linguistic abilities (all *p*s < 0.03). The interaction between temporal damage and NC1 was significant in the full sample (t(27) = 2.09, *p* = 0.046) but not after excluding influential observations (t(24) = 1.43, *p* = 0.16), and the interaction between frontal damage and NC3 (t(27) = 2.09, *p* = 0.046) did not survive outlier exclusion either (*p* = 0.99). Only one interaction was robust to the exclusion of outliers: the interaction between temporal damage and MD network responses during the reading task (sentences > nonwords; full sample: t(27) = 3.10, *p* = 0.004; without influential observations: t(23) = 4.54, *p* < 0.001, n = 29), indicating that in PWA with more extensive temporal damage, stronger MD network responses during reading were associated with better linguistic abilities (**Fig. 6**). No interaction was found for MD responses during the audio-visual task (all *ps* > 0.25). These exploratory findings warrant replication in larger, independent samples with prospective specification of moderation hypotheses.

**Figure 6:**
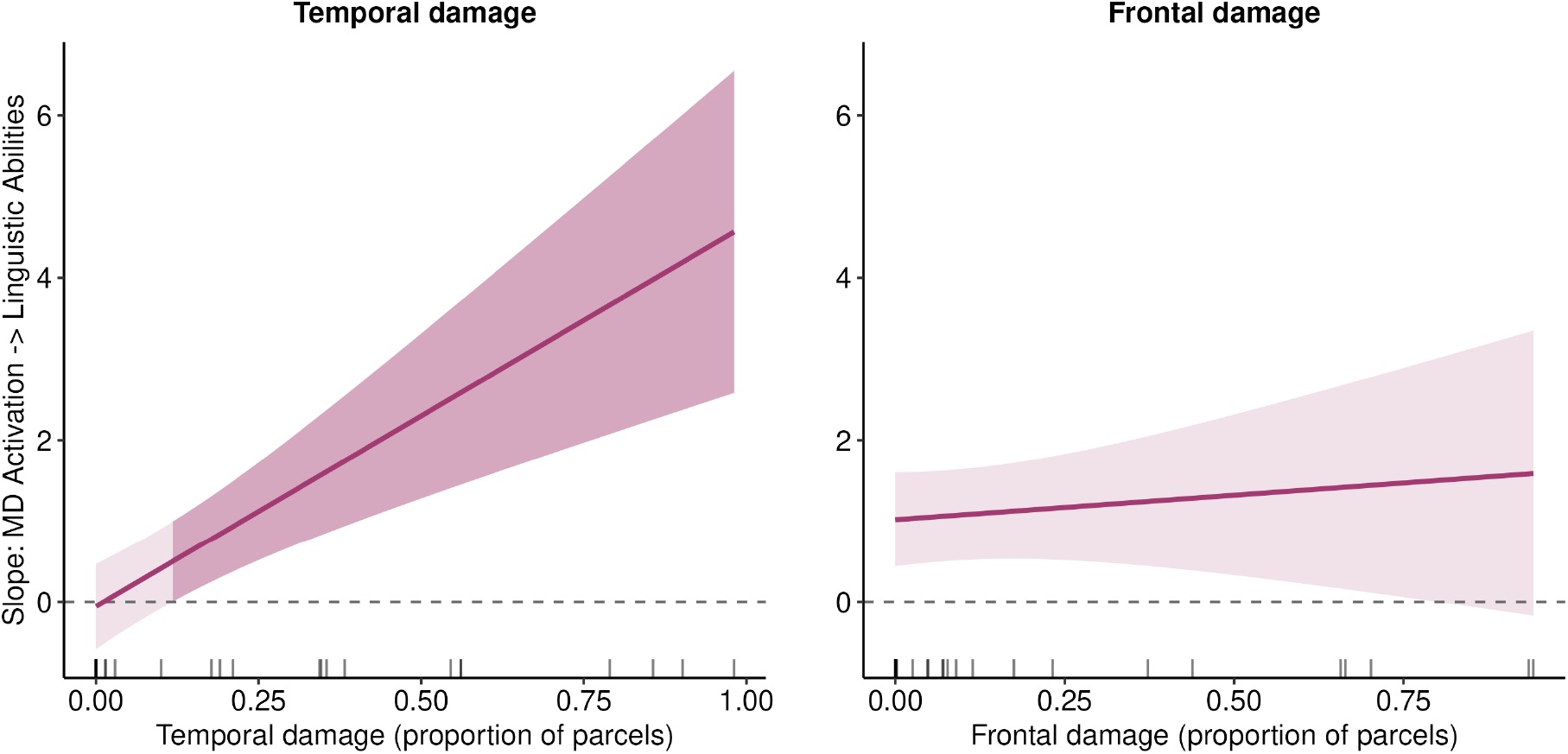
Temporal, but not frontal, stroke damage moderates the relationship between MD network activation during reading and linguistic abilities. Each panel shows the estimated simple slope (± 95% CI) of the sentences > nonwords effect in the multiple demand (MD) network on linguistic abilities (BC1), plotted as a continuous function of the proportion of damage in temporal and frontal language parcels. Slopes were derived from a linear model (BC1 ∼ MD network activation × [Frontal + Temporal damage]; n = 29, after excluding four influential observations with Cook’s distance > 4/n from the initial sample of 33) using the *emtrends* procedure, holding the non-plotted moderator at its sample mean. The dashed horizontal line marks a slope of zero; tick marks along the x-axis indicate each PWA’s observed proportion of damage. The darker shaded region in the left panel marks the range of temporal damage over which the 95% CI excludes zero (i.e., the region of significance), showing that the MD activation–linguistic ability relationship becomes significantly positive only with greater temporal damage (MD activation × Temporal: β = 4.71, t(23) = 4.54, *p* < 0.001). In contrast, the slope did not vary with frontal damage (MD activation × Frontal: β = 0.60, t(23) = 0.65, *p* = 0.52). This analysis is exploratory; results are uncorrected for multiple comparisons and should be interpreted with caution pending replication. **Alt text**: Line graphs of the association between multiple demand network activation and linguistic ability, as a function of lesion extent in language regions. The association strengthens as temporal damage increases. It does not change with frontal damage.

## Discussion

To adjudicate between two accounts of post-stroke language organization—reliance of language processing on the residual portions of the specialized language network versus recruitment of (parts of) the domain-general multiple demand (MD) network for linguistic processing—we carried out a precision fMRI investigation. Using robust and extensively validated fMRI localizers to map the language and MD networks within individuals, we found that in chronic post-stroke aphasia 1) language comprehension relies on the spared parts of the bilateral language network, and 2) the MD network remains functionally dissociated from the language network, showing no language-selective response during language comprehension tasks. Furthermore, 3) better language outcomes are associated with greater engagement of the bilateral language network during language comprehension, not the MD network. Together, these findings suggest that in individuals with chronic post-stroke aphasia, language processing relies on the spared bilateral language regions, whereas the MD regions maintain their role in executive functions and are not recruited for language comprehension more than in age-matched controls.

### Language comprehension relies on the spared bilateral language network in chronic post-stroke aphasia

In individuals with chronic aphasia, the selectivity of the language network is preserved. Spared regions of the bilateral language network showed strong responses to language comprehension across modalities (audio-visual and reading), and little to no response to a non-linguistic demanding task (arithmetic addition), similar to age-matched controls. Both the response magnitude and inter-regional functional connectivity during language comprehension were reduced in the left-hemisphere language network in PWA compared with controls, consistent with a large body of prior work ^48,50,51,53,55–57,155^; see review by ^15^. This activity reduction may reflect physiological factors, such as residual diaschisis in regions connected to the infarcted language areas^156–158^, persistent hypoperfusion around the lesion^159–162^, or vascular changes that lower signal-to-noise ratio^163^. Although most patients in this cohort had mild aphasia (WAB AQ mean = 80.6), it is possible that variable ability to process linguistic stimuli may have influenced the results (**Supplementary Tables 6-7**).

In contrast to the left hemisphere, we found no reliable group differences in magnitude and connectivity in the right-hemisphere language regions in PWA relative to controls, even though it should be noted that PWA show a strong response in all homotopic RH fROIs. Linguistic competence in chronic aphasia was closely linked to how strongly the bilateral language network was engaged, particularly during the reading-based comprehension task. Although we do not find evidence for a right-hemisphere “takeover” of language function beyond activation levels observed in controls, higher right-hemisphere language network activation was consistently related to better linguistic performance across analytic approaches. The association was specific to reading (**Supplementary Table 12**), possibly because reading is more strongly left-lateralized than audio-visual comprehension (**Supplementary Fig. 4**) and therefore more vulnerable to left-hemisphere stroke, making compensatory right-hemisphere engagement particularly beneficial for reading, whereas audio-visual comprehension is already more bilaterally distributed.

Surprisingly, left-hemisphere (LH) activation did not independently predict linguistic outcomes once right-hemisphere (RH) activation and lesion volume were accounted for. However, this pattern was attributable to collinearity between the two hemispheric measures: PWA who more strongly engage residual LH language regions tend to also more strongly engage RH homotopic regions, and the RH feature outperforms the LH feature in predicting better outcomes. This finding suggests that the functional significance of residual LH activation cannot be evaluated in isolation: what matters is not simply whether the damaged left hemisphere remains active, but whether that activity occurs in the context of a synchronously engaged bilateral language network. This construal may help reconcile an apparent contradiction in past studies, where some have reported that RH activation predicts better outcomes in chronic aphasia^60,164^, but others have found that persistent RH reliance is associated with poorer long-term recovery^50,51,55^. The latter likely reflects maladaptive over-recruitment of the right hemisphere in the context of a severely damaged left-hemisphere language network, whereas the former—along with our own findings—likely reflects preserved bilateral network architecture in stroke survivors who retain sufficient LH language resources to sustain coordinated bilateral engagement. Taken together, these results suggest that the right hemisphere’s contribution to language outcomes in chronic aphasia is best understood not as a replacement system, but as a marker of the overall integrity of the bilateral language network.

### The Multiple Demand network maintains a domain-general support role in chronic aphasia

We found no evidence that any parts of the domain-general MD network are recruited by individuals with chronic aphasia to process language. Across our tasks, the MD network in PWA behaved similarly to what has been reported in prior literature for young adults and to what we found in our study for age-matched controls (see also ^44^). In particular, the MD network responded strongly to a cognitively demanding non-linguistic task (arithmetic addition problems), with greater activation for the more difficult problems, and showed little to no response during language processing tasks. For the audio-visual language comprehension task, responses were overall low, close to the fixation baseline; for the reading comprehension task, the MD network of PWA showed a canonical opposite effect, with stronger responses to the meaningless and unstructured nonword-list condition compared to sentences (e.g., ^85,107,108^). Some frontal language regions showed a weak hard > easy arithmetic effect in PWA (LH IFG and RH MFG). However, this effect was also present in controls, often more strongly, and was never larger in PWA, arguing against post-stroke functional reorganization.

Functional connectivity results converged with the activation findings. We found no evidence of consistently increased integration between the language and MD networks during language processing in PWA in either hemisphere. Although the connectivity between the two networks was slightly elevated in the right hemisphere of PWA during rest, this increase was not present during naturalistic story listening, which argues against the idea that the two networks interact to a greater extent in PWA in the service of language comprehension. This dissociation between resting-state and language task-based inter-network connectivity argues against a language-specific reorganization. The MD network therefore appears to retain its domain-general role in supporting cognitively demanding tasks, rather than being repurposed for language comprehension.

PWA showed weaker MD responses to the arithmetic addition task than controls, and this difference was driven by the hard condition. A similar pattern of a selective reduction in the magnitude of response to the hard condition has been reported for healthy older adults compared to younger adults^44^. The aging effect is in line with the Compensation-Related Utilization of Neural Circuits (CRUNCH) hypothesis^165^, whereby the ceiling for the engagement of the executive control network lowers with age. In PWA, however, there are likely additional contributions of direct structural damage to the MD regions or their white-matter connections. Indeed, we found reduced functional connectivity within the left-hemisphere MD network during naturalistic paradigms, including rest, but not within the right-hemisphere MD network, suggesting a stroke-related weakening of ipsilesional MD function that is not limited to tasks with high cognitive demands. Nevertheless, reduced activation for the hard condition was present in MD regions bilaterally, indicating a generalized capacity limitation rather than a purely stroke-related effect.

In line with the lack of engagement of the MD network during language processing, our brain-behavior analyses suggested that properties of the MD network are not reliably associated with linguistic abilities. However, our exploratory analyses did reveal some heterogeneity across patients in the relationship between MD network engagement and language outcomes. In particular, stronger MD responses to the reading comprehension task were associated with better linguistic abilities, but only in patients with extensive damage to the temporal language regions, and not in patients with predominantly frontal damage. Damage to left temporal and parietal cortex has long been shown to lead to longer-lasting and more severe aphasia^151,154^, and some studies have reported qualitatively different functional reorganization patterns following temporo-parietal vs. frontal lesions^55,114^. These preliminary findings are also in line with the Adaptive Plasticity Control (APC) framework ^14^, which proposes that domain-general regions of the MD network may be recruited to scaffold language processing when core language resources are severely compromised, enabling more efficient use of spared linguistic representations and supporting relearning. Our findings suggest that the extent of temporal damage may be a key factor in whether such recruitment occurs. Because these analyses were exploratory and uncorrected for multiple comparisons, future work should replicate them and test the causal contribution of the MD regions to language outcomes. In particular, it should examine whether this contribution changes over the course of stroke recovery and depends on the degree of language network damage. While the results of the present study are preliminary and with a relatively modest sample size of 37 PWA who were generally mild in their aphasia severity, the methodology employed in this work allows a more precise evaluation of the language and the MD networks within each individual.

### Clinical implications and future directions

The current findings carry direct implications for the design of language rehabilitation strategies in aphasia. In particular, rehabilitation efforts should focus on the residual language network given that stronger responses in this network during the chronic stage are associated with better language outcomes. This implication holds for both behavioral interventions, which should focus on linguistic tasks while minimizing general cognitive demands, and for non-invasive brain stimulation. Language localizers can be used to effectively identify the language areas within each patient to guide the placement of stimulating devices. Although some approaches (e.g., transcranial direct current stimulation, tDCS^166,167^) target broad anatomical areas, other, more recent approaches (e.g., focused ultrasound, FUS^168^) can be used to target particular brain areas, which makes individualized functional mapping critical (see Shain et al., under review, for discussion). If the preliminary finding that MD recruitment benefits PWA with extensive damage to the temporal component of the language network replicates in future larger-scale studies, this would suggest that a subset of PWA with severely compromised language resources may benefit from approaches that target executive resources to scaffold linguistic processing. More broadly, our findings align with the general move toward personalized medicine^169,170^ by underscoring the importance of considering individual-level neural and behavioral profiles for both predicting recovery and optimizing treatments^49,171–173^.

### Limitations

Several limitations constrain the interpretation of our findings. **First**, by design, the language tasks used here minimize extraneous cognitive demands and approximate naturalistic comprehension. Although both the critical language conditions in the localizers and the naturalistic story listening task include linguistic features known to increase local processing difficulty, such as low-frequency and unpredictable words and constructions, non-local syntactic dependencies, and temporary ambiguity (e.g., ^103,104^), future studies should manipulate these aspects of linguistic complexity more directly, in both comprehension and production, to test whether the MD network’s engagement may increase with greater linguistic difficulty. This is not the case in young neurotypical adults (see ^106^ for a review), but may be observed in populations with linguistic challenges. **Second**, this study focuses on the chronic stage of recovery with a majority of patients with mild to moderate aphasia; in fact, 15 PWA no longer met the WAB-R criterion for aphasia (AQ ≥ 93.8) at the time of testing, although they all still reported language difficulties affecting their daily life. Whereas most PWA in our sample showed relatively mild comprehension deficits on simple syntactic structures (mean WAB-R auditory verbal comprehension accuracy: 90% ± 14), a majority showed difficulty processing more complex structures (mean accuracy for comprehension of passive sentences and object relative clefts: 65% ± 20). Nevertheless, the lack of a sufficient number of PWA with more severe linguistic impairments may have reduced our ability to detect MD network engagement (if such engagement is only observed in the most impaired PWA). However, two observations mitigate this possibility: aphasia severity did not predict MD network engagement during language processing, and post-hoc analyses restricted to the moderate-to-severe aphasia subgroup yielded the same results (**Supplementary Table 10**). Furthermore, we cannot exclude the possibility that the MD network plays a role during earlier stages of recovery, before language network function normalizes^55^. Indeed, some prior evidence supports a time-limited scaffolding role for the MD network: for example, fronto-parietal functional connectivity during the subacute stage predicts subsequent language recovery^115^, and MD activity during speech production at the acute stage predicts long-term outcomes^65^. Longitudinal studies that track the engagement of and functional connectivity in the language and MD networks from the acute through the chronic stages of recovery will be critical to understanding the potential changes in the MD network’s contributions to language processing in aphasia.

**Third**, the study has a sex imbalance between the groups: PWA were predominantly male (30 men, 7 women; 19% female), whereas the control group was more balanced (17 men, 21 women; 55% female). This asymmetry reflects the higher incidence of stroke in men in the relevant age range, but potentially complicates the interpretation of group differences in the degree of language lateralization and interhemispheric connectivity, although large-scale datasets of the language responses suggest no reliable differences between sexes (e.g., ^174^; unpublished data from the Fedorenko lab).

## Conclusion

Using precision fMRI, the present study shows that people with chronic post-stroke aphasia do not recruit the domain-general multiple demand (MD) network to a greater extent than neurologically healthy age-matched individuals during language comprehension. Instead, similar to control participants, language comprehension in people with aphasia relies on the spared parts of the bilateral language network. Despite weaker responses and lower within-network functional connectivity following a stroke, the language-selective and the domain-general MD networks, identified with robust individual-level fMRI localizers, remain dissociated. The approach used here to identify the spared parts of the language network within individuals is straightforwardly portable to the clinic and can be used to identify the most promising targets for non-invasive brain stimulation to enhance language recovery and improve outcomes for individuals with aphasia.

## Data Availability

The data that support the findings of this study will be available on ResearchBox.

## Acknowledgements

We thank all individuals who participated in this study and their caregivers. We additionally acknowledge present and past members of the Boston University Center for Brain Recovery and the Fedorenko Lab at MIT. We acknowledge the Athinoula A. Martinos Imaging Center at the McGovern Institute for Brain Research, MIT, and the Cognitive Neuroimaging Center at Boston University. For technical support during scanning, we thank Steve Shannon and Atsushi Takahashi at MIT, and Shruthi Chakrapani and Stephanie McMains at BU. We also thank Hope Kean and Kris Brewer for their help in creating the audio-visual version of the language localizer.

## Funding

This work was supported by the NIH - National Institute on Deafness and Other Communication Disorders (grant R01DC016950 to SK and EF). EF was additionally supported by research funds from the McGovern Institute for Brain Research, MIT’s Simons Center for the Social Brain, MIT’s Poitras Center for Psychiatric Disorders Research, and the MIT Siegel Family Quest for Intelligence.

## Competing interests

Swathi Kiran is an advisor and owns stock in Constant Therapy Health, a digital therapy company, but there is no scientific overlap with the work reported in this paper. The other authors report no competing interests.

## Notes

### Author Declarations

All participants gave written informed consent in accordance with the requirements of MIT Committee on the Use of Humans as Experimental Subjects (COUHES) and the Institutional Review Board of Boston University. The Massachusetts Institute of Technology Committee on the Use of Humans as Experimental Subjects (COUHES) and the Institutional Review Board (IRB) of Boston University approved the conduct of the study.

### Summary of Updates

This version of the manuscript has been revised to update the following: (1) the statistical models, which now include by-participant random slopes, with corrections for multiple comparisons applied consistently across analyses; (2) the brain-behavior analyses, restricted to participants whose behavioral testing was completed within 9 months of scanning (n = 33), with an updated neural principal component analysis, elastic net results and partial regression plots (Fig. 5); (3) new lateralization and spatial-overlap results; (4) a more complete description of the behavioral assessments and of the exploratory lesion-location analyses; and (5) corresponding revisions to the Results, Discussion, figures and supplementary material. The main conclusions are unchanged.

